# Altered brain dynamics across bipolar disorder and schizophrenia revealed by overlapping brain states

**DOI:** 10.1101/2022.10.07.22280835

**Authors:** Jean Ye, Huili Sun, Siyuan Gao, Javid Dadashkarimi, Matthew Rosenblatt, Raimundo X. Rodriguez, Saloni Mehta, Rongtao Jiang, Stephanie Noble, Margaret L. Westwater, Dustin Scheinost

## Abstract

Aberrant brain dynamics putatively characterize bipolar disorder (BD) and schizophrenia (SCZ). Previous studies often adopted a state discretization approach when investigating how individuals recruited recurring brain states. Since multiple brain states are likely engaged simultaneously at any given moment, focusing on the dominant state can obscure changes in less prominent but critical brain states in clinical populations. To address this limitation, we introduced a novel framework to simultaneously assess brain state engagement for multiple brain states, and we examined how brain state engagement differs in patients with BD or SCZ compared to healthy controls (HC). Using task-based data from the Human Connectome Project, we applied nonlinear manifold learning and K-means clustering to identify four recurring brain states. We then examined how the engagement and transition variability of these four states differed between patients with BD, SCZ, and HC across two other international, open-source datasets. Comparing these measures across groups revealed significantly altered state transition variability, but not engagement, across all four states in individuals with BD and SCZ during both resting-state and task-based fMRI. In our post hoc and exploratory analysis, we also observed associations between state transition variability and age as well as avolition. Our results suggest that disrupted state transition variability affects multiple brain states in BD and SCZ. By studying several brain states simultaneously, our framework more comprehensively reveals how brain dynamics differ across individuals and in psychiatric disorders.

## Introduction

Bipolar disorder (BD) and schizophrenia (SCZ) are multifaceted, heterogeneous psychiatric disorders characterized by psychotic symptoms and impaired executive functioning (Murray et al., 2004; Eisenberg & Berman, 2010; Cotrena et al., 2020). These illnesses and their cardinal symptoms putatively reflect disruptions in the interactions between functional brain networks (Chase & Phillips, 2016; Sheffield & Barch, 2016; Dong et al., 2018). Many studies of functional networks in these illnesses have studied these disruptions from the perspective of “static” functional network structure estimated over the course of a minutes-long scan. Nevertheless, as communications within the brain are dynamic and fluctuate over small time windows (Hutchison et al., 2013; Lurie et al., 2020), our understanding of the neural correlates underpinning these disorders may be incomplete.

Indeed, a new wave of methods have emerged to capture this additional temporal information in the form of brain states, or distinct, recurring patterns of brain activity or connectivity (Lurie et al., 2020). Brain states can be characterized in terms of state engagement (i.e., the proportion of time points spent in a particular state, also called dwell time) and transition variability (i.e., the number of times individuals transitioned from one state to another, also known as transition). These methods have revealed aberrant brain dynamics in BD and SCZ that were undetectable using static approaches (Rashid et al., 2016; Du et al., 2017; Nguyen et al., 2017). Compared to healthy controls (HC), individuals with these disorders demonstrate aberrant dwell time in brain states and altered transitions between them (Damaraju et al., 2014; Du et al., 2016; Reinen et al., 2018; Miller et al., 2016; Rashid et al., 2014; Du et al., 2021; Wang et al., 2021; Wang et al., 2022). Brain dynamic alterations, in turn, are associated with psychiatric symptoms in these groups, including elevated suicide risk, psychotic symptom severity, and hallucinations (Wang et al., 2022; Wang et al., 2021; Miller et al., 2016; Reinen et al., 2018).

However, these studies mainly assigned each time unit to only one state despite the possibility that the activity or connectivity patterns in a given time window can be considered a combination of several patterns (Miller et al., 2016). Individuals are also likely to recruit multiple states simultaneously to support many complex behaviors, such as cognitive control. Thus, this state discretization approach might ignore meaningful but non-dominant states, and potentially misrepresent dynamic information. For instance, assigning each time window to only one state makes it challenging to evaluate whether the elevated recruitment of one state might accompany reduced engagement of another as an alternative or compensatory mechanism in clinical populations.

To address this limitation, we developed a novel method that combines nonlinear manifold learning and non-negative least squares regression to simultaneously assess frame-to-frame state engagement for multiple states. We applied 2-step Diffusion Mapping (2sDM) and nonlinear manifold learning to project fMRI data from the Human Connectome Project (HCP; Van Essen et al., 2013) onto a low-dimensional space, and we subsequently identified reproducible brain activity patterns across multiple fMRI tasks (Gao et al., 2021). Next, we used non-negative least squares regression to extend these recurring brain activity patterns to two international datasets to examine how individuals with BD, SCZ, and HC engaged and transitioned through these brain states while at rest. We hypothesized that individuals with BD and SCZ would show aberrant overall state engagement and transition variability compared to HC during rest. Since difficulties with executive functioning and cognitive flexibility have been consistently reported in individuals with BD and SCZ (O’Donnell et al., 2016; Cotrena et al., 2016; Eisenberg & Berman, 2010; Orellana & Slachevsky, 2013), secondary analyses were performed using fMRI data from a task-switching paradigm to further investigate state transition in clinical populations and link altered brain state dynamics to clinical symptoms. We found aberrant state transition variability in clinical populations and evidence suggesting that decreased state transition variability was associated with avolition symptoms and older age. Our approach simultaneously provides individualized, moment-to-moment information for multiple states, painting a more complete picture of brain dynamic alterations in patients with BD and SCZ.

## Methods

### Participants

Data from three independent, publicly available datasets were analyzed in this study. First, we used the HCP S500 release (Van Essen et al., 2013) to identify recurring brain states. The moment-to-moment engagement of the brain states identified in HCP was explored in HC, BD, and SCZ groups from the UCLA Consortium for Neuropsychiatric Phenomics (CNP) and the University of Tokyo Hospital site in the Japanese Strategic Research Program for the Promotion of Brain Sciences (SRPBS) datasets (Poldrack et al., 2016; Tanaka et al., 2021). Patient diagnoses were made by trained clinicians using DSM-IV criteria for BD and SCZ (Poldrack et al., 2016; Tanaka et al., 2021). For the SRPBS dataset, the HC group was screened using the Mini-International Neuropsychiatric Interview. HCs with lifetime psychiatric disorder diagnoses were excluded from the CNP dataset. Demographic information for each dataset is shown in Table 1 (see **Supplementary Table 1** for patient participants’ medication information).

**Table 1.** Demographic information by dataset

| SRPBS |  |  |  |
| --- | --- | --- | --- |
|  | HC | BD | SCZ |
| N | 86 | 40 | 33 |
| Self-reported sex (F/M) | 56/30 | 15/25 | 11/22 |
| Age (years) (M±SD) | 47.07±15.18 | 34.48±9.11 | 31.64±10.77 |
| CNP (Resting-state dataset) |  |  |  |
|  | HC | BD | SCZ |
| N | 103 | 35 | 38 |
| Self-reported sex (F/M) | 49/54 | 15/20 | 9/29 |
| Age (years) (M±SD) | 30.29±7.99 | 34.09±9.01 | 35.42±8.99 |
| CNP (task-switching dataset) |  |  |  |
|  | HC | BD | SCZ |
| N | 120 | 49 | 48 |
| Self-reported sex (F/M) | 56/64 | 21/28 | 11/37 |
| Age (years) (M±SD) | 31.65±8.84 | 35.29±9.03 | 36.19±8.87 |

HC: healthy control; BD: bipolar disorder; SCZ: schizophrenia

HCP fMRI data from six different tasks (motor, working memory, social, emotional, relational and gambling) were used to identify recurring brain states. As the HCP dataset contains tasks ranging from simple motor to complex cognitive and affective paradigms, it has the potential to reveal brain states underlying a rich set of cognitive processes. Additionally, finding brain states in an independent dataset allows us to avoid circular analysis and overfitting when examining them in the CNP and SRPBS cohorts. We used the resting-state fMRI data from CNP and SRPBS dataset for primary analyses. Task-based fMRI data from the CNP task-switching paradigm were used for secondary analyses.

### FMRI data preprocessing

Acquisition and imaging parameters for the HCP, CNP and SRPBS datasets have been detailed elsewhere (Van Essen et al., 2013; Poldrack et al., 2016; Tanaka et al., 2021). The HCP minimal preprocessing pipeline was used for the HCP dataset (Glasser et al., 2013). Standard preprocessing procedures described in previous work (Gao et al., 2021) were applied to the structural and functional data from both the CNP and SRPBS datasets. Briefly, we removed the first four volumes to allow magnetic field stabilization for the SRPBS dataset. After brain extraction with OptiBet (Lutkenhoff et al., 2014), structural data was nonlinearly registered to the standard MNI-152 space. For functional data, slice time and motion correction were performed using SPM8 before being linearly aligned to the structural data. Data cleaning for all datasets was performed with BioImage Suite. Regression of covariates of no interest, including linear and quadratic drift, a 24-parameter model of motion, and mean white matter, cerebrospinal fluid and gray matter signals, was performed. Timeseries data were temporally smoothed (cutoff frequency approximately ∼0.12Hz) and then extracted using the Shen-268 atlas (Shen et al., 2013) from all resting state fMRI datasets for each participant. We applied the same preprocessing pipeline and quality control procedures to the task-based fMRI data used in our secondary analysis. Several brain nodes were excluded from resting-state (n=7) and task-based (n=5) brain dynamics analyses due to noise and/or incomplete coverage. For the CNP and SRPBS data, we additionally scrubbed time points with over 0.45 framewise displacement, and we excluded participants with over 20% of their time points censored due to motion.

Additional quality control criteria for the HCP and CNP datasets were described in a previous study (Gao et al., 2021). One CNP participant was excluded due to an incomplete resting-state scan. Fourteen SRPBS participants were excluded from further analysis after not meeting preprocessing quality control benchmarks (i.e., issues with skull stripping, nonlinear or linear registration). After these exclusions, 390 HCP participants remained for brain state identification. For brain dynamics analyses, 336 participants from the CNP and SRPBS datasets were included in the resting-state analysis (177 participants from the CNP dataset, HC: n=104, BD: n=35, SCZ: n=38; 159 participants from the SRPBS dataset: HC: n=86, BD: n=40, SCZ: n=33, **Table 1**), and 217 participants (CNP dataset, HC: n=120, BD: n=49, SCZ: n=48; **Table 1**) were included in the task-based analysis.

### Brain state identification

As detailed information on 2sDM was described in an earlier work (Gao et al., 2021), we provide a brief overview here. Nonlinear approaches like 2sDM can project complex neural data onto a manifold that is more representative of its temporal dynamics than linear methods to find robust brain states (Gao et al., 2021). In brief, diffusion maps was applied twice to fMRI timeseries data: first, to reduce fMRI timeseries in the participant’s and brain node’s dimensions, and second, to embed it into a lower-dimensional space, where time points showing similar activity were located close to each other (**Figure 1**). We applied K-means clustering for 100 iterations using the first three embedding dimensions and found the optimal number of clusters, or brain states, as four using the Calinski-Harabasz criterion (Caliñski & Harabasz, 1974). These labels can be applied back to the original data to find time points that are associated with each state across participants. For each brain state, all associated fMRI data from all participants were averaged first across the individual level and next across time points to establish a representative timepoint.

**Figure 1:**
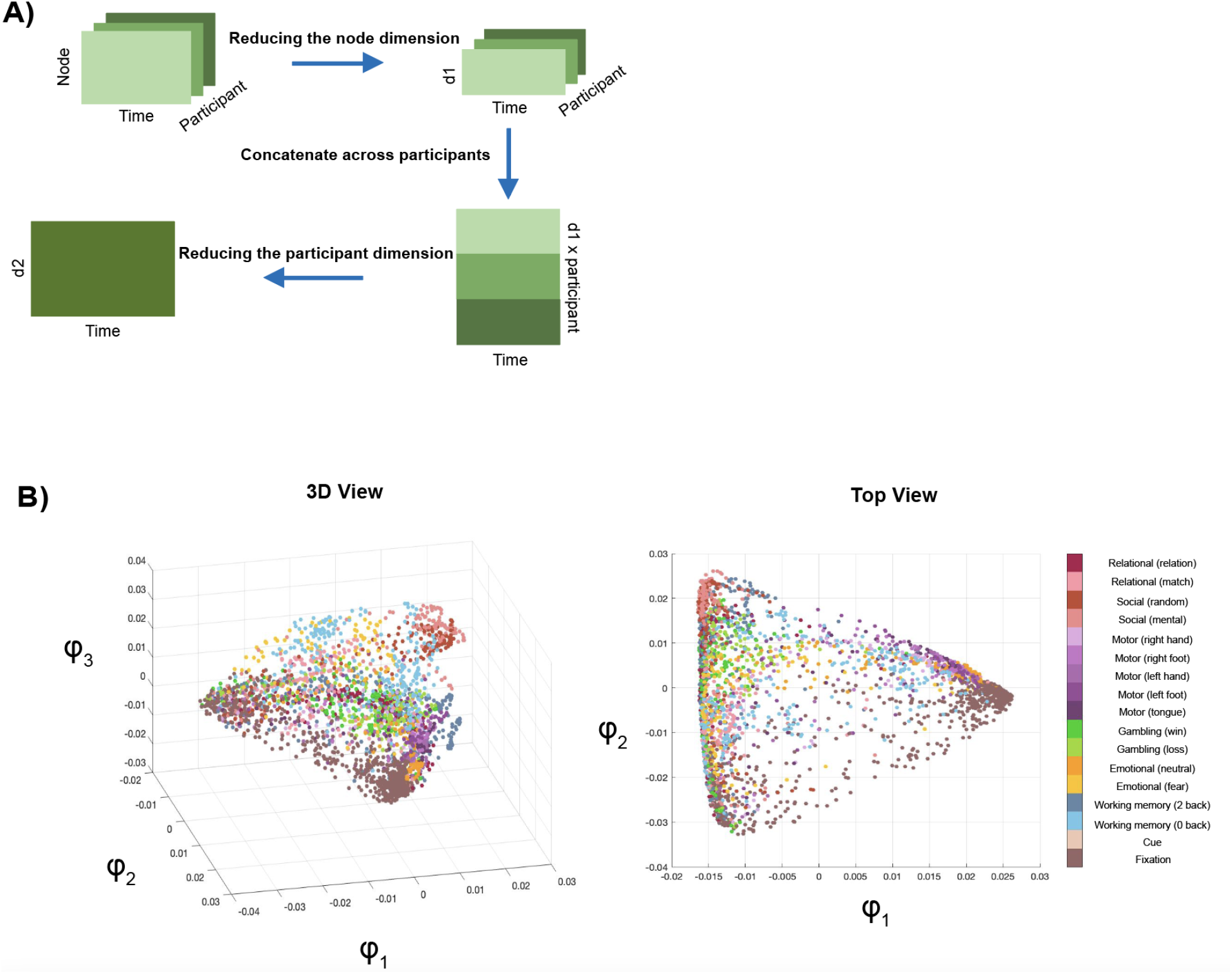
2sDM pipeline. **A)** is a flowchart showing how 2sDM was applied to the HCP dataset to first reduce the brain node’s (d1) and participant’s (d2) dimensions before projecting it to a low-dimensional space. **B)** provides several views of the manifold. Each dot here represents a time point, color-coded by the task condition it is associated with. Further details about 2sDM can be found in Gao et al., 2021.

To understand which canonical functional networks were associated with each brain state, we first identified the activated and deactivated regions within each representative time point (defined as having activation above or below 0, respectively). Next, we extracted the activation and deactivation percentages of each network using the ten functional networks previously defined with the Shen-268 atlas (Finn et al., 2015; Noble et al., 2017). Percentages were computed by dividing the number of activated or deactivated regions by the total number of regions in a network for each representative time point. This procedure allowed us to account for the different network sizes and to see how different networks activate or deactivate for each brain state.

### Resting-state brain dynamics analyses

To evaluate moment-to-moment brain state engagement, we regressed each representative time point from every CNP and SRPBS rest time point using non-negative least squares regression (implemented with the MATLAB function *lsqnonneg;* **Figure 2**). Non-negative least squares was selected over standard regression to ensure brain state engagements were positive, as a negative state engagement, which is possible with standard regression, might be difficult to interpret (Lee & Seung, 1999). It fits with the intuition that multiple brain states might be engaged simultaneously at any given moment, allowing us to study their overlapping, moment-to-moment contribution. Representative time points and all rest time points were first normalized by standard deviation before regression.

**Figure 2:**
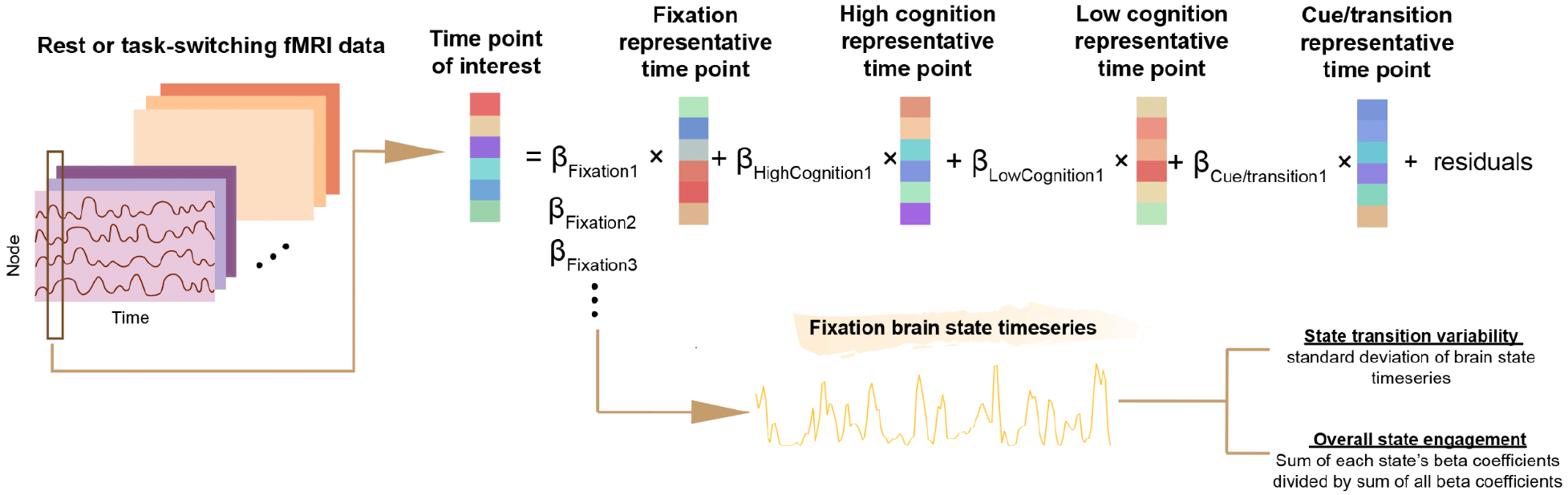
Assessing moment-to-moment brain state engagement with non-negative least squares regression. We first identified four recurring brain states using the HCP dataset and characterized them as fixation, high cognition, low cognition and cue/transition. The representative time points for each state were next regressed from each resting-state or task-switching time point of interest. This flowchart shows an example of how beta coefficients for the fixation state can be concatenated across time to create a brain state timeseries. The two extracted summary measures, state transition variability and overall state engagement, are the main brain dynamic measures focused in this study.

By concatenating the output beta coefficients for each state across time, we can create a brain state timeseries indicating a particular state’s weighted contribution over time for each participant (**Figure 2**). Two summary measures can be extracted from each brain state timeseries: 1) overall state engagement —a continuous version of state engagement computed by dividing the sum of each state’s beta coefficients by the sum of all beta coefficients, and 2) state transition variability —a continuous version of state transition defined as the standard deviation of brain state timeseries.

### Secondary analysis with task-based fMRI data

As our resting-state results suggested aberrant state transition variability in clinical populations, we opted to study altered state transition further by performing secondary analyses in a smaller sample with task-based fMRI data. This task-switching paradigm contained 96 total trials and required participants to flexibly switch between task rules (i.e., responding to the color vs. the shape of the stimulus) based on the cue presented (Poldrack et al., 2016). Overall state engagement and transition variability were extracted and studied in this dataset.

### Statistical analysis

We combined the two datasets for group comparison to maximize statistical power for detecting brain dynamic alterations in BD and SCZ and to examine the extent to which these alterations generalize across cultures. Both datasets collected eye-open resting-state scans from adult participants (Poldrack et al., 2016; Tanaka et al., 2021). As we have an overall state engagement and state transition variability measure for each of the four brain states, multivariate group differences were examined across all three groups using MANCOVA, with site, medication, age and sex as covariates.The group comparison analysis was then supplemented by assessing group differences between both patient groups and HC (i.e., BD vs HC and SCZ vs HC) with t-squared test, including site, medication, age and sex as covariates.

Cohen’s d effect sizes in pairwise differences were computed to quantify how much each brain state contributed to multivariate group differences (Bettinardi, 2022). For our primary and secondary analyses, p<0.0125 (i.e., p<0.05/4) was considered significant after Bonferroni correction for multiple comparisons. We did not correct for multiple comparisons for our post hoc or exploratory analyses.

### Comparison with a state discretization approach

We additionally examined brain dynamics using a state discretization approach to more directly evaluate our results in the context of previous studies. For this analysis, each rest and task time point was first correlated with each state representative time point and then assigned to the state showing the highest correlation value. State transition was measured as the number of times there was a change in state assignment from one time point to the next, whereas dwell time was operationalized as the number of time points assigned to one state divided by the total number of time points. As the state transition measure here required a continuous assessment, we did not censor based on motion. But participants with excessive motion (criteria same as described above) were excluded from this analysis. We then compared dwell time and state transition across groups using ANCOVA, with site (when appropriate), medication, age and sex as covariates. ANCOVA was used here since the dwell time measures were not completely independent from each other, and there was only one state transition measure.

### Exploratory analysis with behavioral data

To further understand the implications of differences in state transition variability, we examined the association between overall state transition variability and symptom measures in patient participants. We focused on brain dynamics measures from task-based fMRI data, as the sample size was larger here relative to the resting-state dataset. To estimate overall state transition variability, we applied principal component analysis (PCA) to the state transition variability measures from all brain states using data from the clinical groups. The first component scores were then correlated with measures from the Scale for Assessment of Negative Symptoms (Andreasen, 1983) and Scale for Assessment of Positive Symptoms (Andreasen, 1984) in the BD and SCZ groups using Spearman correlation. We specifically focused on avolition, anhedonia, and attention since the other factor measures were zero inflated (**Supplementary Figure 1**). These symptom measures were only available in clinical populations not in HCs. As similar state transition variability patterns were found in both clinical groups during task, data from participants with BD and SCZ were combined in this analysis to increase statistical power and to examine transdiagnostic associations between altered brain dynamics and clinical symptoms.

## Results

### Brain states and canonical functional brain networks

Replicating previous work (Gao et al., 2021), we identified four brain states with distinct activity patterns using 2sDM and K-means clustering. We then characterized these brain states as high cognition, low cognition, fixation and cue/transition based on the task conditions involved in each cluster (**Figure 3A**) and examined the extent to which each canonical network contributed to each state’s representative time point.

**Figure 3:**
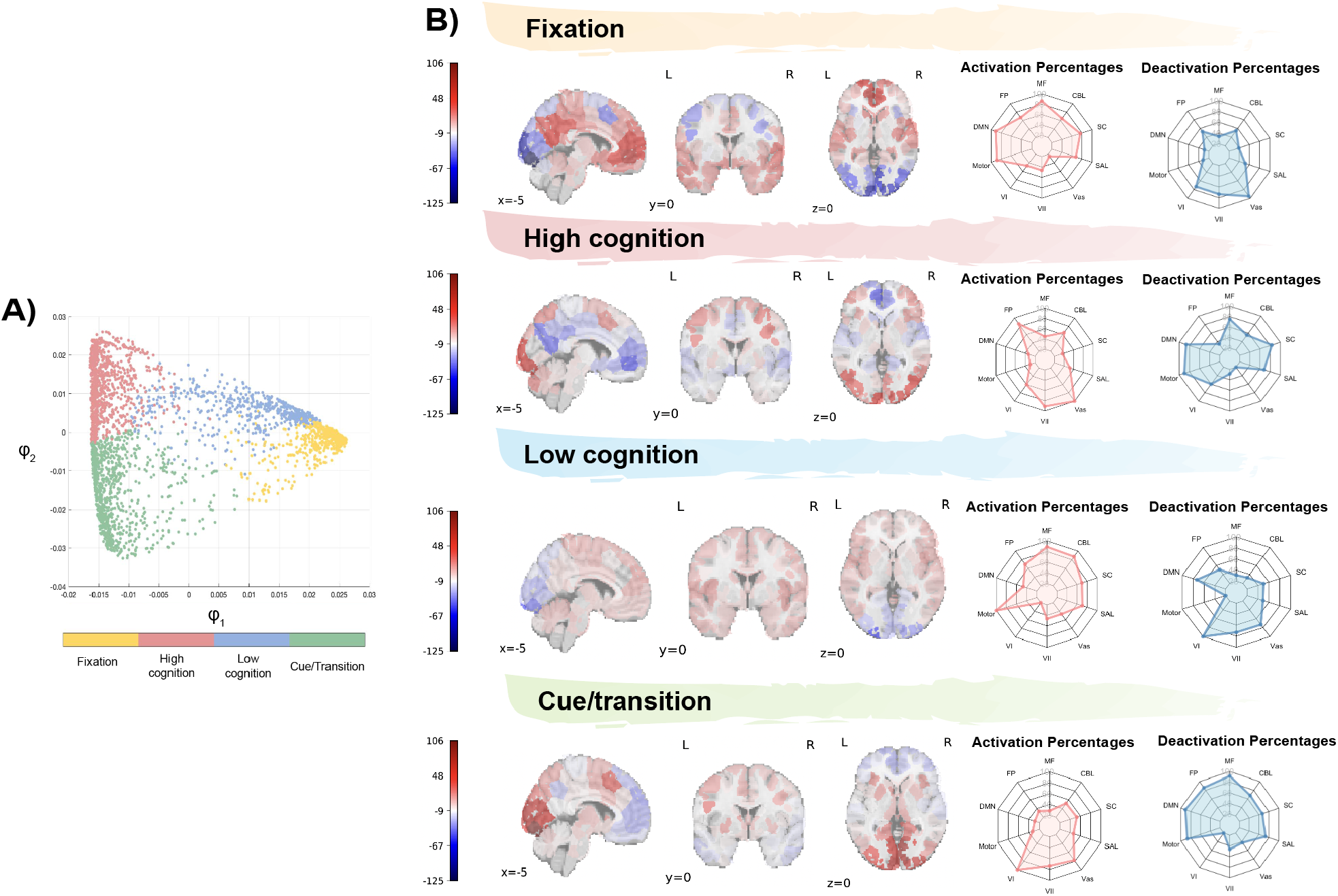
Recurring brain states and their association with canonical functional brain networks. **A)** indicates the four brain state output from K-means clustering. These states were characterized as fixation, high cognition, low cognition, and cue/transition based on the prominent task conditions associated with the time points in each cluster (each dot here represents a time point; see **Figure 1** for their task conditions). **B)** visualizes the four representative time points. Spider plots show the relative extent to which functional brain networks were activated and deactivated in each representative timepoint. For illustrative purposes, the raw activation values were multiplied by 100. MF, medial frontal network; CBL, cerebellum network; SC, subcortical network; SAL, salience network; Vas, visual association network; VII, visual II network; VI, visual I network; motor, motor network; DMN, default mode network; FP, frontoparietal network.

We found that default mode network (DMN), motor network, and medial frontal network (MF) had the highest activation percentage in the fixation brain state (DMN: 88.89%; Motor: 85.71%; MF: 82.76%; **Figure 3B**), while the visual networks had the highest deactivation percentage (visual association (VAs): 94.44%; Visual I: 66.67%; Visual II: 66.67%; **Figure 3B**). For high cognition, the VAs network, the Visual II network and the frontoparietal (FP) network had the highest activation percentage (VAs: 100%; Visual II: 88.87%; FP: 82.35%; **Figure 3B**), whereas the motor network, the DMN and the subcortical network had the highest deactivation percentage (Motor: 87.76%; DMN: 83.33%; subcortical: 79.31%; **Figure 3B**). The motor network, the medial frontal (MF) network and the cerebellum network had the highest activation percentage in the low cognition brain state (Motor: 100%; MF: 86.21%; cerebellum: 84%; **Figure 3B**), while the DMN and the visual networks had higher deactivation percentage for this state (Visual I: 100%; Visual II: 66.67%; VAs: 66.67%; DMN: 66.67%; **Figure 3B**).

All three visual networks showed high activation percentage for the cue/transition brain state (Visual I: 100%; VAs: 72.22%; Visual II: 66.67%; **Figure 3B**), but the MF network, the DMN and the motor network showed the highest deactivation percentage (MF: 89.66%; DMN: 88.87%; Motor: 83.67%; **Figure 3B**). In general, the networks associated with each brain state align with the presumed cognitive processes invoked by each state. For example, the entire motor network was activated for the low cognition state, which contained time points from the motor task. Additionally, the high cognition state, which included time points from working memory, emotional, social and relational tasks, was associated with FP activation and DMN deactivation.

### Primary and secondary analysis: Resting-state and task brain dynamics across groups

Before group comparisons, we first examined the residual term from non-negative least squares regression. We did not find any rest or task time point with an outlier residual term (defined as 1.5 times the interquartile range), suggesting that the activity patterns of all rest and task time points can be considered as some weighted combinations of the four brain states obtained from the HCP.

During rest, we observed a significant main effect of diagnosis group on state transition variability (MANCOVA; F(8,648) =3.093, p=0.002; **Figure 4B**) but not for overall state engagement (MANCOVA; F(8,648)=1.327, p=0.227; **Figure 4A**). There were additionally main effects of site and age on state transition variability (**Supplementary Table 2**). Main effects of medication use were not significant (**Supplementary Table 2**).

**Figure 4:**
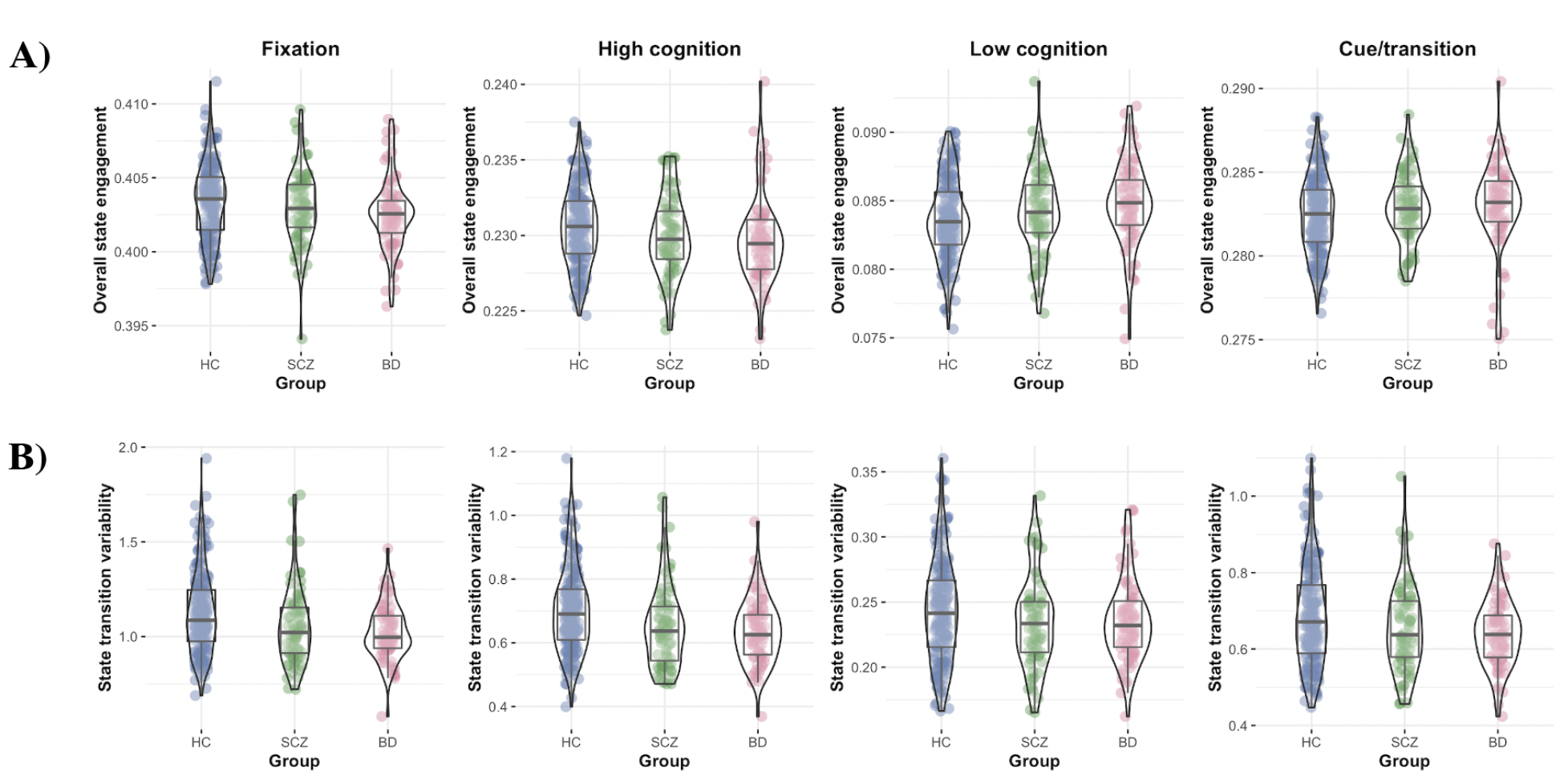
Brain dynamics during rest. **A)** All three groups showed similar overall state engagement across all four states during rest. **B)** But we observed lower state transition variability across all four states in BD and SCZ compared to HC. Blue, HC; green: SCZ; red: BD.

Follow-up tests were performed to understand how each clinical group differed from HC for state transition variability. Participants with BD (t-squared; F(4,253)=4.850, p<0.001) and SCZ (t-squared; F(4,249)=2.883, p=0.023) showed significant differences in state transition variability compared to HCs when including site, medication, age, and sex as covariates (**Supplementary Table 3**). Both clinical groups showed lower state transition variability across all four states, with the high cognition brain state showing the largest effect size (**Supplementary Table 4**).

During task, we similarly observed significant main effects of diagnostic group on state transition variability (MANCOVA; F(8,412)=2.536, p=0.011; **Figure 5B**) but not on overall state engagement (MANCOVA; F(8,412)=0.734, p=0.662; **Figure 5A**). While the main effect of medication was not significant, there were significant main effects of age, site, and sex (**Supplementary Table 5**). With medication, age, and sex as covariates, both participants with SCZ (t-squared; F(4,157)=3.103, p=0.017) and BD (t-squared; F(4,158)=2.792, p=0.028) demonstrated lower state transition variability across most states compared to HCs (**Supplementary Table 6**), with the high cognition state again showing a large effect size (**Supplementary Table 7**).

**Figure 5:**
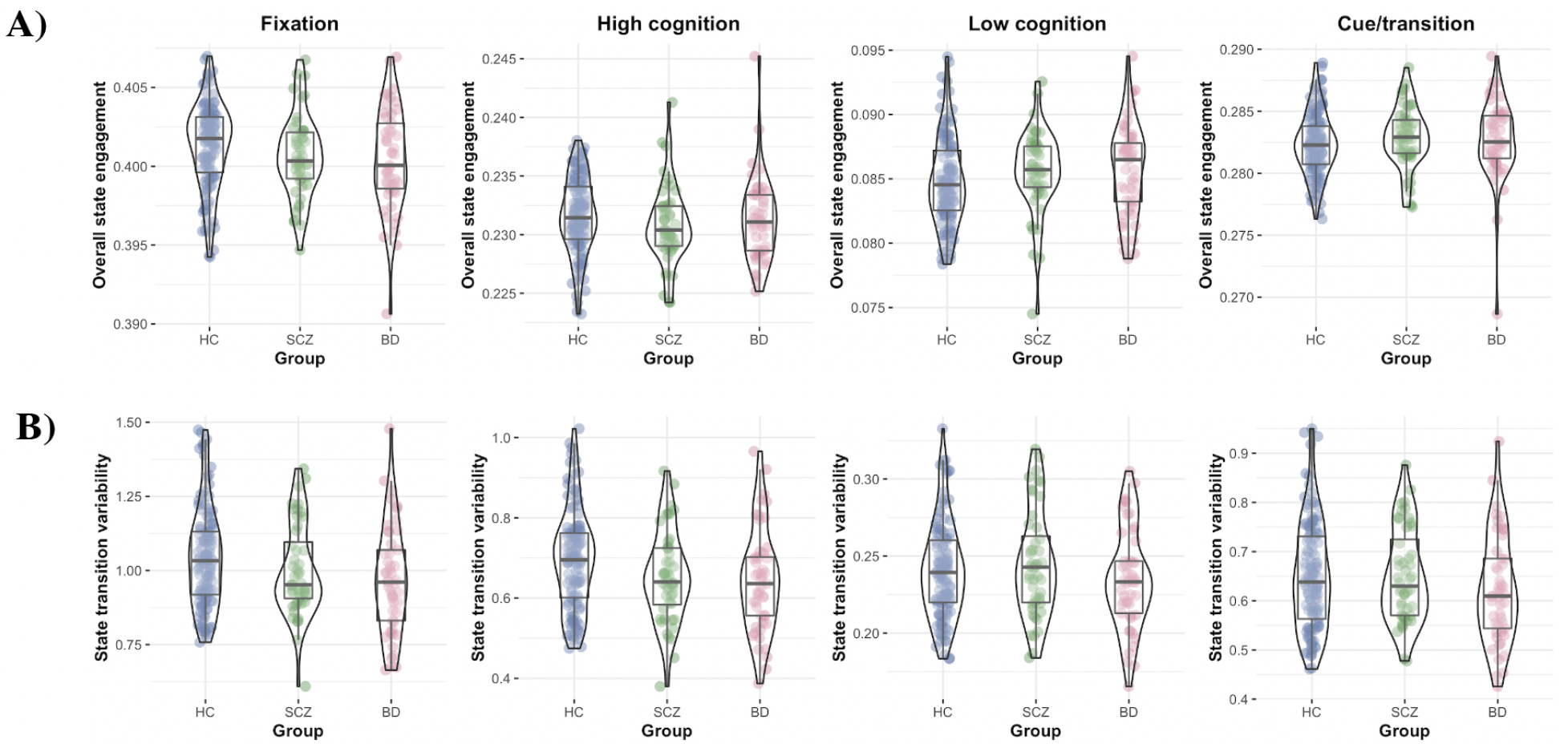
Brain dynamics during task-switching. **A)** Similar to rest, we found that all three groups recruited the four brain states similarly during task-switching. **B)** But clinical populations showed significantly different state transition variability compared to HC. While BD showed lower state transition variability across all four states, SCZ showed decreased fixation and high cognition but elevated low cognition and cue/transition state transition variability. Blue, HC; green: SCZ; red: BD.

### Brain dynamics with a state discretization

With discretized states, we did not observe a significant group effect on state transition during rest (F(2,326)=1.404, p=0.247; **Supplementary Table 8**) or task-based runs (F(2,208)=0.167, p=0.846; **Supplementary Table 9**). Similarly, the main effect of diagnostic group on dwell times was not significant for both rest and task data following Bonferroni correction (**Supplementary Table 8 and 9**).

### Post hoc analysis examining the association between age and state transition variability

To further understand the significant main age effect, we correlated age with overall state transition variability within each group during rest and task in a post hoc analysis. Overall state transition variability was estimated by applying PCA to the four state transition variability measures. The 1st component accounted for 94.424% of the variance in rest and 92.526% in task. During rest, older HCs showed lower state transition variability (r=-0.414, p=2.854×10^×9^; **Figure 6A**). While there was also a significant negative correlation in SCZ (r=-0.348, p=0.003; **Figure 6A**), this effect was smaller in BD (r=-0.192, p=0.098; **Figure 6A**). The association between age and state transition variability was significant in all three groups during the task (HC: r=-0.269, p=0.003; BD: r=-0.355, p=0.012; SCZ: r=-0.318, p=0.028; **Figure 6B**).

**Figure 6:**
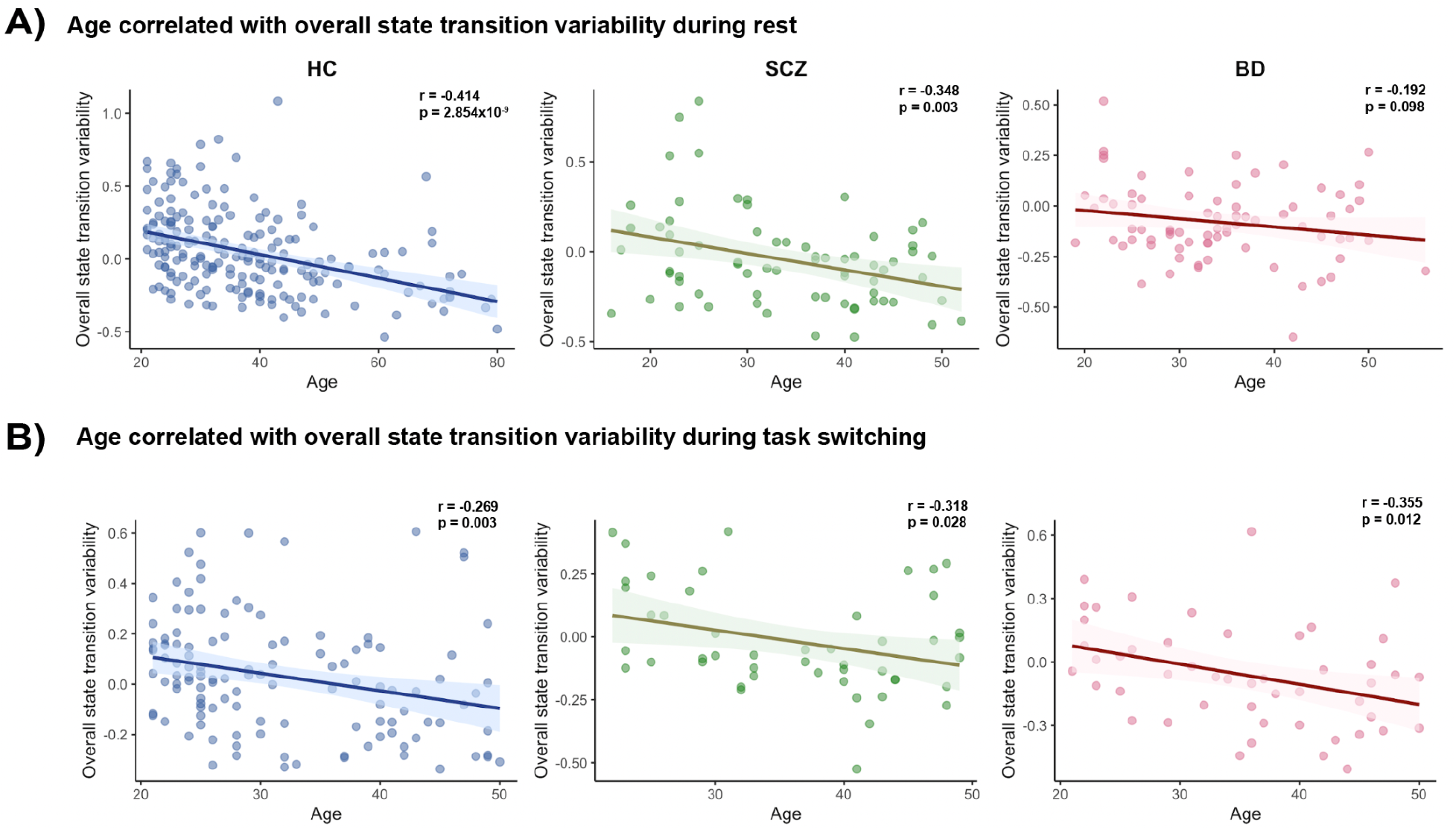
Associations between state transition variability and age. **A)** We observed a significantly negative correlation between overall state transition variability and age in HC and SCZ during rest, but not in BD. **B)** In task-switching, older participants showed decreased overall state transition variability across all three groups.

### Exploratory analysis studying the associations between altered state transition variability and symptom measure

Using data from the two clinical groups, the first PCA component on overall state transition variability accounted for 92.56% of the variance. Our post hoc analysis revealed that lower overall state transition variability during task-switching was associated with elevated avolition in individuals with BD and SCZ (r=-0.222, p=0.029; **Figure 7**), but not the other symptom measures (attention: r=0.154, p=0.132; anhedonia: r=-0.152, p=0.138; **Figure 7**).

**Figure 7:**
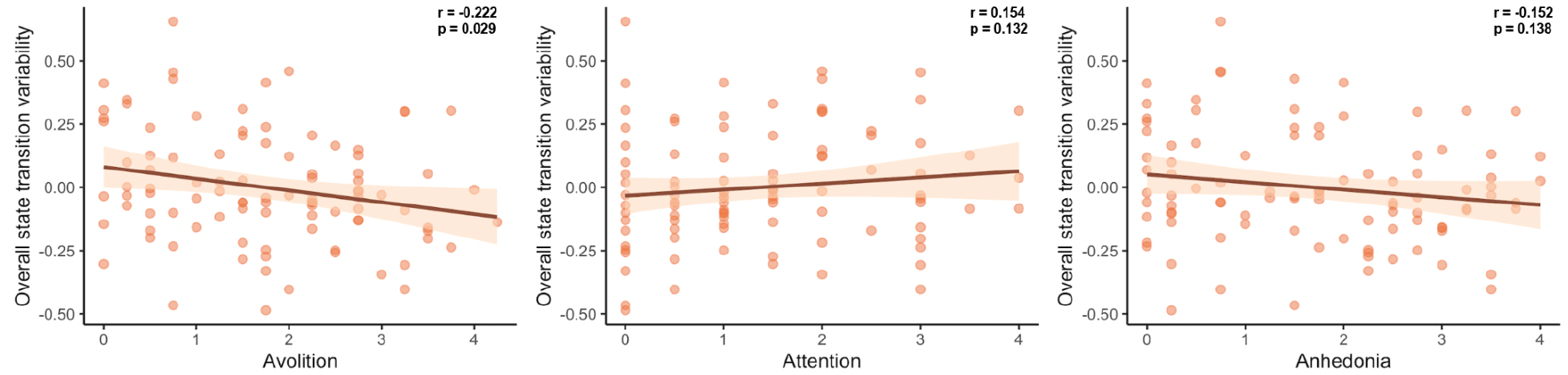
Associations between state transition variability and clinical symptoms. Elevated avolition symptoms were associated with decreased overall state transition variability. But state transition variability was not correlated with attention or anhedonia.

## Discussion

Here, we leveraged nonlinear manifold learning and non-negative least squares regression to assess the continuous engagement of multiple brain states simultaneously during resting-state and task-based neuroimaging. After deriving four brain states from the HCP dataset, we detected aberrant state transition variability across these states in two independent datasets of individuals with BD and SCZ. Both clinical groups showed reduced state transition variability across all four states during rest. While the same alteration patterns were observed in most states for task, we also found a pattern of higher low cognition and cue/transition state transition variability in the SCZ group. However, our version of state discretization approach did not reveal aberrant state transition in clinical populations, indicating that our framework might be more sensitive to altered brain dynamics than a state discretization approach. Strong associations between overall state transition variability and age were observed. These results deepen our understanding of the neural correlates of BD and SCZ and highlight the importance of considering multiple brain states simultaneously when studying psychiatric disorders.

### Aligning our work with previous studies using a state discretization approach

Other works examining brain dynamics in BD and SCZ have found mixed results, reporting increased, similar, and decreased state transition and dwell time in clinical populations (Damaraju et al., 2014; Rashid et al., 2014; Du et al., 2016; Miller et al., 2016; Du et al., 2021; Wang et al., 2021). Varying methods might have contributed to this discrepancy. However, our work suggests that aberrant state transition variability is a global deficit affecting all brain states in BD and SCZ, potentially complementing previous findings. Since our approach allows us to examine multiple states concurrently, we found that altered state transition can manifest in various ways for different states (i.e., decreased or increased variability). This was shown in our task results in the SCZ group. It can potentially explain the mixed findings regarding state transition in clinical populations, as one can come to very different conclusions depending on which state is considered as dominant and, therefore, highlighted in a state discretization approach. Notably, our version of the forced state approach did not elucidate significant group differences in state transition, suggesting that the current approach might be more powerful in detecting brain dynamic alterations.

We found similar overall state engagement across patient and control groups even though previous studies have revealed altered dwell time in BD and SCZ (Damaraju et al., 2014; Du et al., 2016; Reinen et al., 2018; Wang et al., 2021). One possible explanation is that our overall state engagement measure might not be a direct equivalent of dwell time. A state with less dwell time (i.e., considered dominant in fewer time points) can still show high overall state engagement if it is consistently engaged in the background. However, our results also suggest that a state discretization approach might mask variability differences as dwell time differences. For instance, if a state demonstrated decreased transition variability (as in most states examined here), it would likely show consistent engagement across time and be assigned to more time points if it is a dominant state or fewer if it is less prominent. This difference in variability could then be interpreted as increased or decreased dwell time, respectively. Our results suggest that state transition variability, rather than dwell time or overall state engagement, was the main factor driving brain dynamic differences in the clinical populations examined here.

### Linking the dopaminergic system with brain dynamic differences

The dopaminergic system is a possible convergent mechanism for the various topics discussed in this paper, including SCZ (Maia & Frank, 2016; Howes & Kapur, 2009, Tost et al., 2010), BD (Cousins et al., 2009), cognitive flexibility (Cools & D’Esposito, 2011), aging (Bäckman et al., 2006; Li et al., 2009), and avolition (Heinz et al., 1998). Emerging works further suggest that changes in the dopaminergic system can lead to brain dynamic differences. A recent network control theory study (Braun et al., 2021) found that dopamine gene expression and pharmacological manipulations of dopaminergic tone were associated with changes in state transition and stability. Altered brain dynamics in SCZ was also reported in that study (Braun et al., 2021). Decreased dopamine has additionally been associated with changes in executive functions as people gradually age (Bäckman et al., 2000; Erixon-Linroth et al., 2005). One natural speculation following previous findings in aging and clinical populations is that fluctuations in dopamine neurotransmission can affect the coordination between brain states to influence behaviors.

### Studying brain dynamic changes in other critical developmental periods

This study and our earlier discussion focused on adults, but other critical developmental periods should also be examined further. Adolescence, a period of dramatic brain changes (Yurgelun-Todd, 2007; Blakemore, 2008; Sturman & Moghaddam, 2011), has been associated with the emergency of psychiatric disorders (Belfer, 2008; Costello et al., 2011) and a more active dopaminergic system (Wahlstrom et al., 2010). Previous work investigating brain dynamics in this population has additionally reported a positive association between age and brain state transition (Medaglia et al., 2018), the opposite of that in older adults (Ezaki et al., 2018; Lee et al., 2022). Systematically investigating the trajectory of brain dynamic changes across the lifespan can allow the identification of markers indicating the early emergence of psychiatric disorders and provide insight into whether individuals with these markers might experience more drastic behavioral and brain dynamic alterations as they age.

### Detecting altered brain dynamics with task-based fMRI data

Here, we supplemented our resting-state analysis with fMRI data that were collected during performance of a cognitive flexibility task. Compared to the HC group, individuals with BD and SCZ demonstrated aberrant transition variability across states during task switching. This might suggest that altered brain dynamics underlie the difficulties with executive functioning often reported by these patients (O’Donnell et al., 2017; Contrena et al., 2016; Eisenberg & Berman, 2010; Orellana & Slachevsky, 2013). Furthermore, altered brain dynamics were detected in a smaller sample (n=217) using a fMRI task paradigm that specifically challenged the ability to respond flexibly to changing cognitive demands. This result supports prior arguments that more robust brain-behavior relationships can be identified when the construct of interest (cognitive flexibility in this case) is engaged during the scan (Greene et al., 2018; Finn, 2021).

### Limitations and future directions

Our approach allows a more complete picture of brain dynamics by tracking multiple states at the same time, and it can be flexibly applied to a large variety of datasets. For example, future work can apply this method to fMRI data that are acquired during naturalistic paradigms, which can advance understanding of how brain state engagement fluctuates to support cognition in a more ecologically valid task. Additionally, we use multivariate analysis to examine activation at the whole-brain level, which has increased statistical power compared to univariate analysis (Noble et al., 2022). Studying the brain at this broader level can potentially better reveal the neural mechanisms underpinning complex psychiatric disorders. While many studies have turned to time-varying functional connectivity to study dynamic changes in whole-brain patterns, most of these approaches require a few time points to measure the correlation between brain regions’ activity levels. It can then be challenging to extract moment-specific information. As we rely on brain activation during tasks to identify brain states and measure their engagement, we were able to provide frame-to-frame state engagement information and additionally had some insight into the cognitive processes each brain state might be associated with. But as recent approaches have demonstrated the feasibility of studying the interaction between brain regions on a more frame-to-frame basis (Esfahlani et al., 2022), future work should more systematically compare various brain dynamic approaches. These evaluations can inform whether different methods provide complementary information or if one method might be preferable in certain scenarios.

A few limitations of the present study bear further investigation. As we used open-source data, some clinical information, such as medication information, was unavailable for the SRPBS participants included in this study. However, when including medication as a covariate for our task-based analyses in the CNP dataset, we still observed aberrant state transition variability in patients with either BD or SCZ, which would suggest that group differences are potentially robust to medication effects. Future work should investigate how medication type, dose, and treatment history relate to brain dynamics in these patient groups. Since we did not group participants with BD based on their mood state at the time of the scan, future research should examine whether euthymic, manic or depressive mood states might be associated with variations in brain dynamics. Finally, only a limited number of participants were included in our analysis studying the relationship between altered brain dynamics and clinical symptoms. Although we increased our sample size by combining both clinical groups, future work should explore the association between state transition variability and symptoms in a larger sample.

## Conclusion

In summary, we introduced a novel approach to investigate multiple brain states that co-occur simultaneously, which painted a more comprehensive picture of brain dynamics in a population-based cohort, as well as in patients with serious mental illness. Our results suggest that aberrant state transition variability—instead of overall state engagement—drives brain dynamic differences in BD and SCZ. We also observed a robust association between state transition variability and age. As less dominant brain states are often neglected when states are discretized, our work demonstrates that approaches assessing multiple brain states simultaneously can better reflect the inherent complexity of brain dynamics.

## Data Availability

The HCP dataset is available at https://db.humanconnectome.org/. The CNP dataset can be accessed through https://openneuro.org/datasets/ds000030/. The SPRBS dataset can be obtained through https://bicr-resource.atr.jp/srpbsopen/.

## Acknowledgements

Data was provided in part by the Human Connectome Project, WU-Minn Consortium (Principal Investigators: David Van Essen and Kamil Ugurbil; 1U54MH091657 funded by the 16 NIH institutes and Centers that support the NIH Blueprint for Neuroscience Research; and by the McDonnell Center for Systems Neuroscience at Washington University) and the Consortium for Neuropsychiatric Phenomics (NIH Roadmap for Medical Research grants UL1-DE019580, RL1MH083268, RL1MH083269, RL1DA024853, RL1MH083270, RL1LM009833, PL1MH083271, and PL1NS062410). The CNP dataset was accessed through OpenNEURO (https://openneuro.org/datasets/ds000030/). A portion of the data used in the preparation of this work were obtained from the DecNef Project Brain Data Repository (https://bicr-resource.atr.jp/srpbsopen/), collected as part of the Japanese Strategic Research Program for the Promotion of Brain Sciences (SRPBS) supported by the Japanese Advanced Research and Development Programs for Medical Innovation (AMED). JY was supported by the Gruber Science Fellowship from the Gruber Foundation through Yale University. MR was supported by the National Science Foundation Graduate Research Fellowship under Grant No. DGE-2139841. RXR was supported by the National Research Service Award (award number: 5T32GM100884-09) from the National Institute of General Medicine. SN was supported by the National Institute of Mental Health under award number K00MH122372. MLW was supported by the National Institute on Drug Abuse (T32DA022975). DS was supported by R01MH121095. We thank Ignacio Ruiz-Sanchez, Huda Siddiqui and Iris Cheng for data quality control help.

## Supplementary materials

**Supplementary Table 1.** Medication information for the CNP dataset

| CNP (Resting-state dataset) |  |  |
| --- | --- | --- |
|  | BD | SCZ |
| Antipsychotic | 8 | 28 |
| Antidepressant | 5 | 3 |
| Mood stabilizer | 13 | 2 |
| Others | 2 | 1 |
| CNP (Task-switching dataset) |  |  |
|  | BD | SCZ |
| Antipsychotic | 11 | 33 |
| Antidepressant | 6 | 6 |
| Mood stabilizer | 18 | 4 |
| Others | 4 | 1 |

**Supplementary Figure 1.**
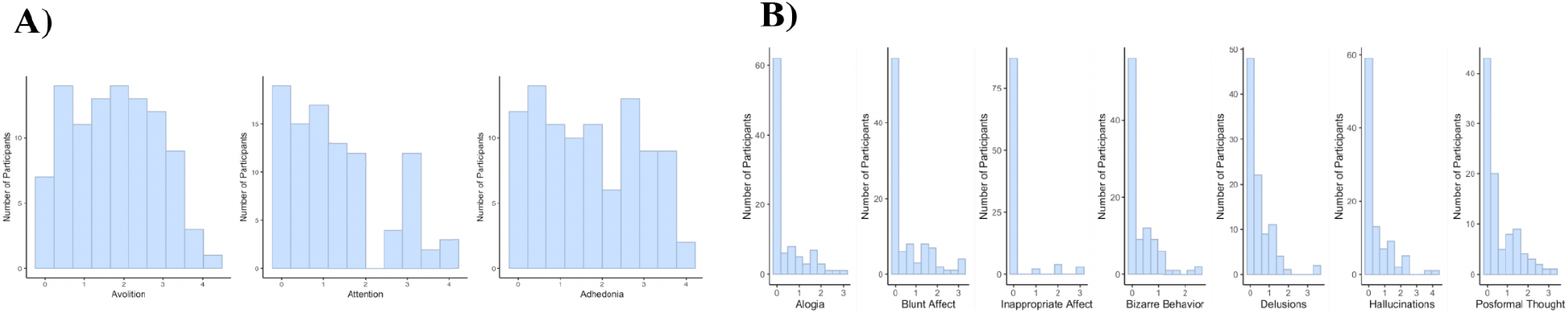
Distributions of clinical symptom scores. **A)** showed the distribution of symptom scores used to correlate with state transition variability. We did not perform further analysis with the symptom scores shown in **B)** since the distribution was zero-inflated.

**Supplementary Table 2.** Resting-state MANCOVA covariates

| State transition variability |  |  |
| --- | --- | --- |
|  | F-stat | p |
| Site | F(4,323)=23.884 | $2.2 \times 10^{-16}$ |
| Medication | F(16,1304)=1.078 | 0.371 |
| Age | F(4,323)=8.749 | $1.024 \times 10^{-6}$ |
| Sex | F(4,323)=1.337 | 0.256 |
| Overall state engagement |  |  |
| Site | F(4,323)=3.734 | 0.005 |
| Medication | F(16,1304)=1.584 | 0.066 |
| Age | F(4,323)=1.793 | 0.130 |
| Sex | F(4,323)=0.857 | 0.490 |

**Supplementary Table 3.** Resting-state state transition variability T-squared covariates

| HC vs. BD |  |  |
| --- | --- | --- |
|  | F-stat | p |
| Site | F(4,253)=20.803 | $7.72 \times 10^{-15}$ |
| Medication | F(16,1024)=1.232 | 0.236 |
| Age | F(4,253)=6.070 | 0.0001 |
| Sex | F(4,253)=1.745 | 0.141 |
| HC vs. SCZ |  |  |
| Site | F(4,249)=18.753 | $1.72 \times 10^{-13}$ |
| Medication | F(16,1008)=1.264 | 0.213 |
| Age | F(4,249)=6.084 | 0.0001 |
| Sex | F(4,249)=1.130 | 0.343 |

**Supplementary Table 4.** Resting-state state transition variability effect sizes

| HC vs. BD |  |
| --- | --- |
| State | Cohen's d |
| Fixation | 0.499 |
| High cognition | 0.524 |
| Low cognition | 0.213 |
| Cue/transition | 0.394 |
| HC vs. SCZ |  |
| State | Cohen's d |
| Fixation | 0.317 |
| High cognition | 0.382 |
| Low cognition | 0.233 |
| Cue/transition | 0.272 |

**Supplementary Table 5.** Task-switching MANCOVA covariates

| State transition variability |  |  |
| --- | --- | --- |
|  | F-stat | p |
| Medication | F(16,832)=1.195 | 0.265 |
| Age | F(4,205)=9.352 | 5.81x10<sup>-7</sup> |
| Sex | F(4,205)=3.727 | 0.006 |
| Overall state engagement |  |  |
| Medication | F(16,832)=1.642 | 0.053 |
| Age | F(4,205)=1.876 | 0.116 |
| Sex | F(4,205)=1.424 | 0.227 |

**Supplementary Table 6.**
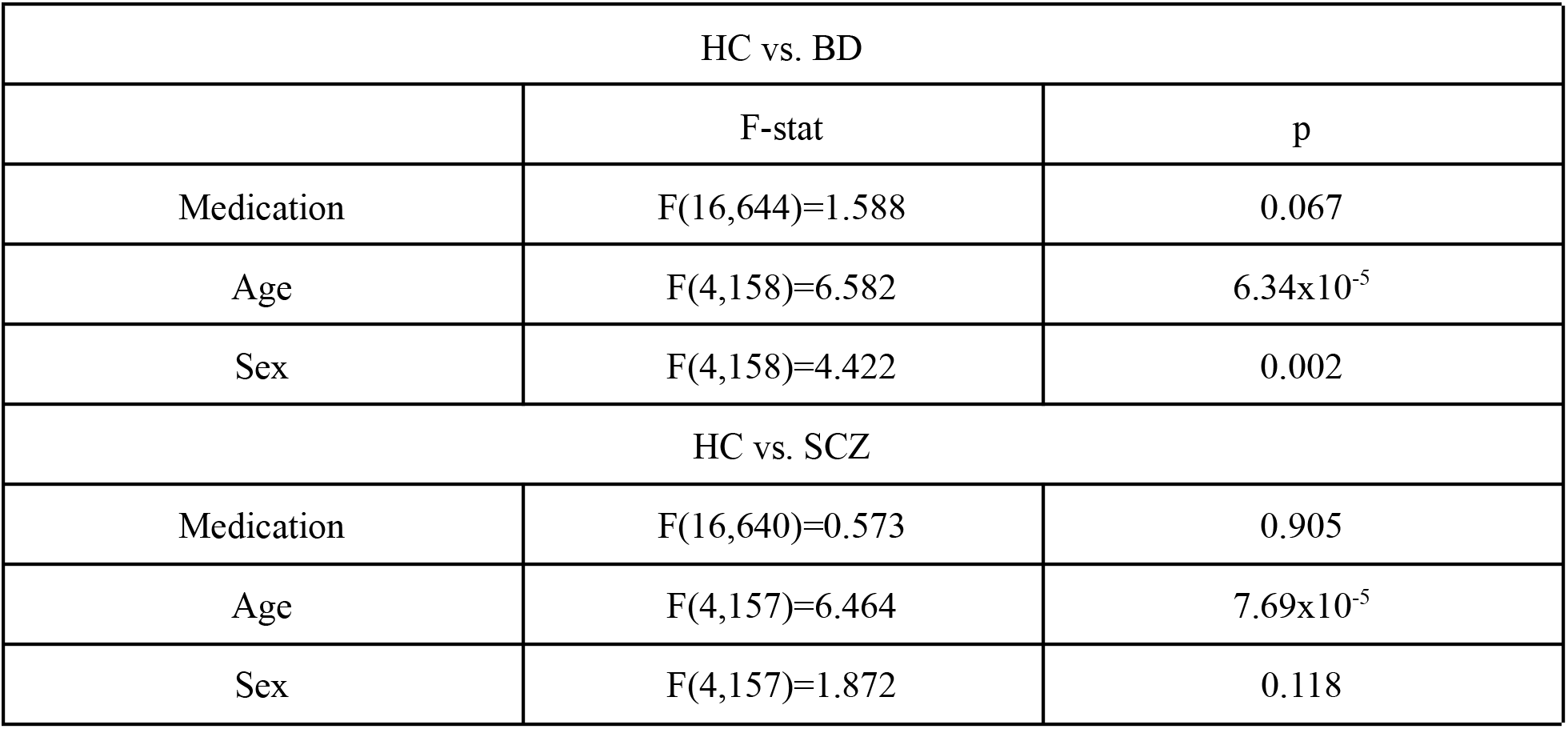
Task-switching state transition variability T-squared covariates

**Supplementary Table 7.** Task-switching state transition variability effect size

| HC vs. BD |  |
| --- | --- |
| State | Cohen's d |
| Fixation | 0.420 |
| High cognition | 0.415 |
| Low cognition | 0.201 |
| Cue/transition | 0.246 |
| HC vs. SCZ |  |
| State | Cohen's d |
| Fixation | 0.245 |
| High cognition | 0.349 |
| Low cognition | -0.129 |
| Cue/transition | -0.011 |

**Supplementary Table 8.**
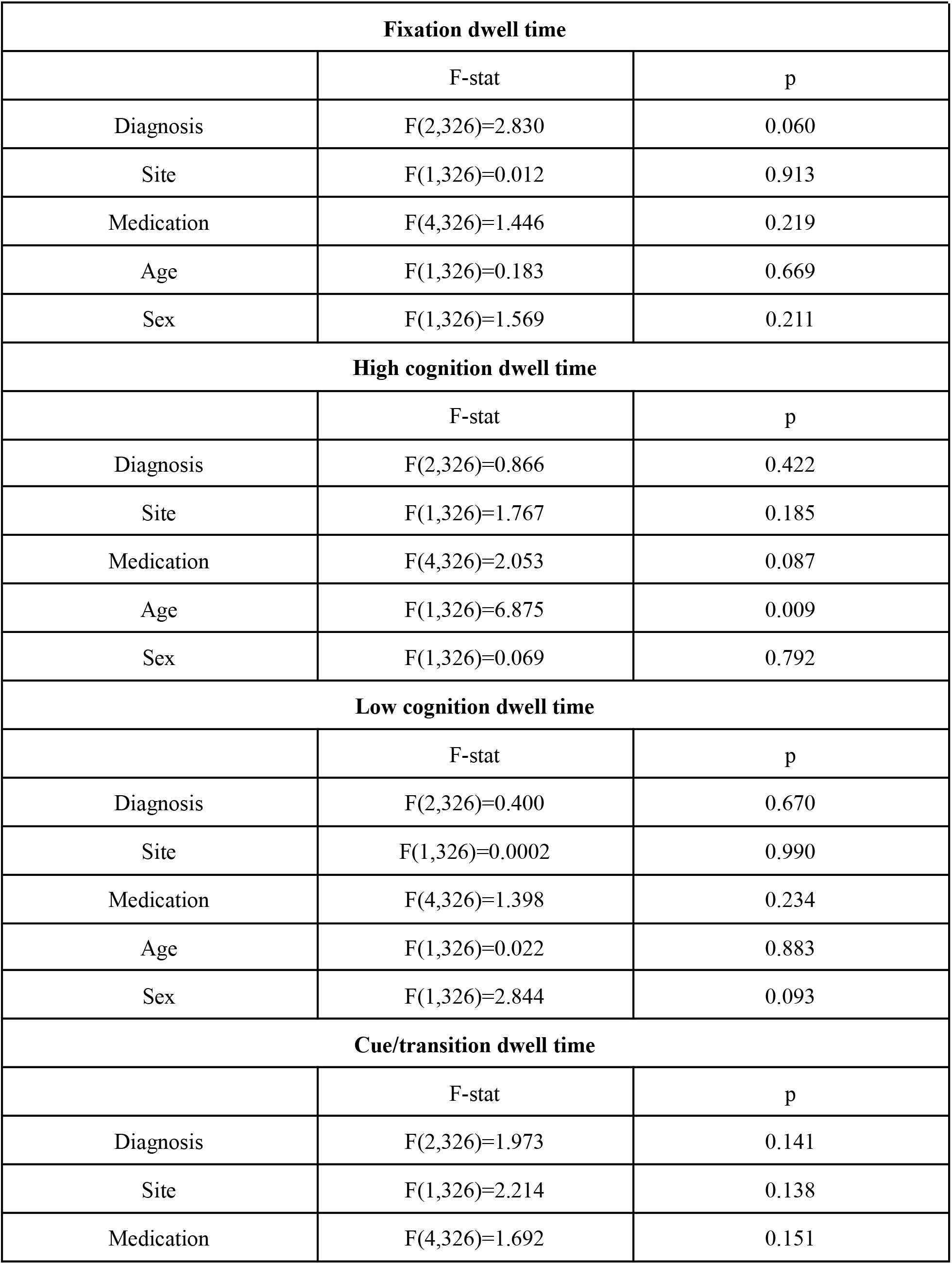
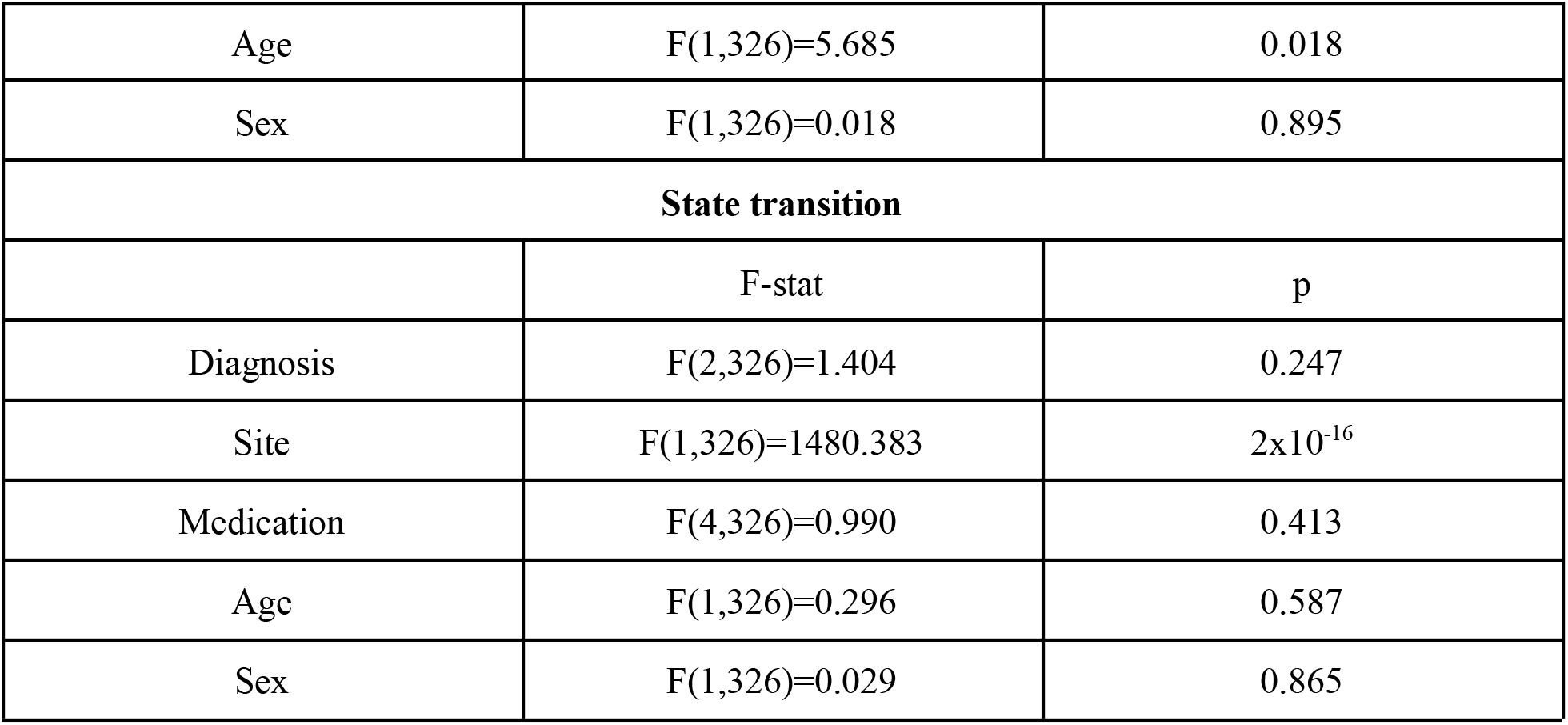
Discrete state approach results for resting-state data

**Supplementary Table 9.**
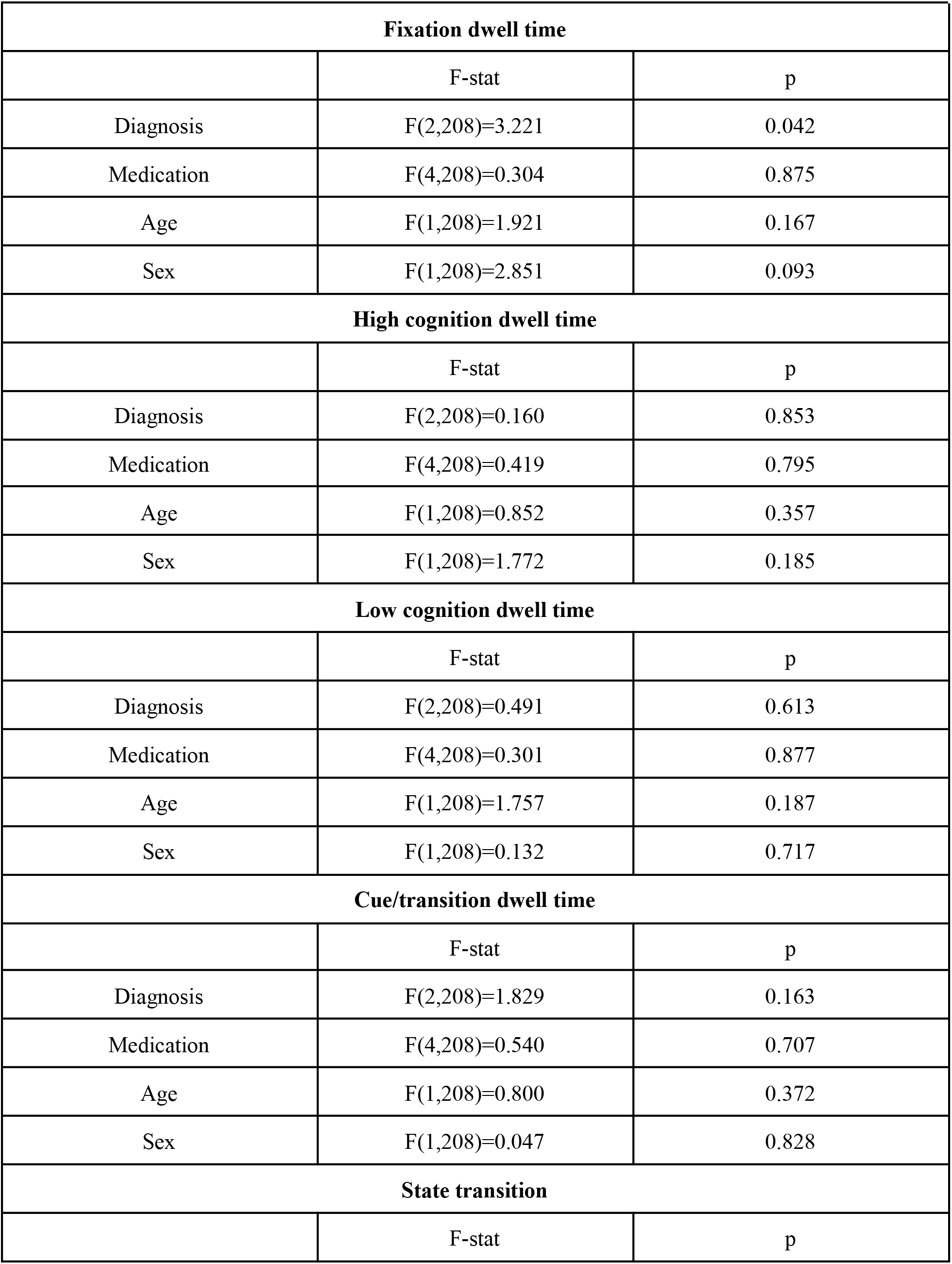
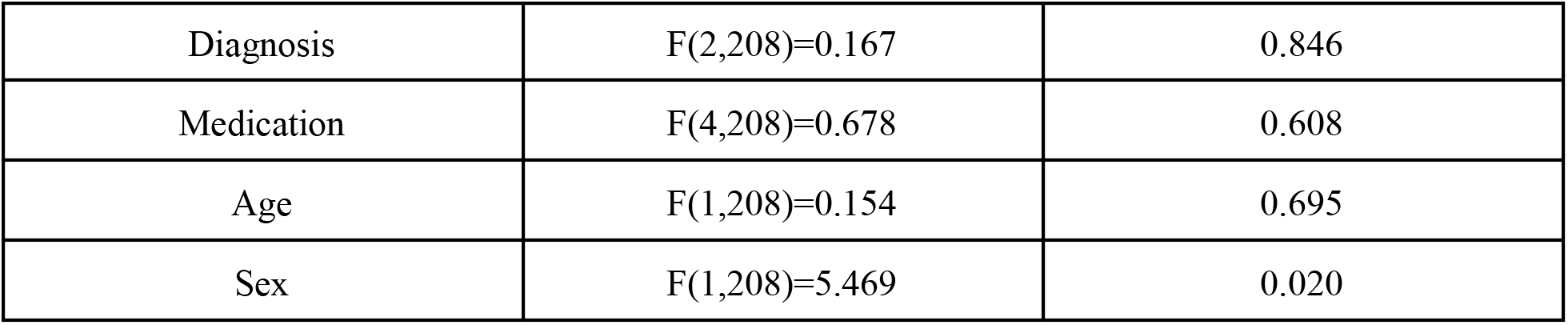
Discrete state approach results for task-switching data

